# Paternal body mass index before conception associated with offspring’s birth weight in Chinese population: a prospective study

**DOI:** 10.1101/2021.06.17.21258438

**Authors:** Renying Xu, Weixiu Zhao, Tao Tan, Haojie Li, Yanping Wan

## Abstract

Whether paternal epigenetic information of nutrition might be inherited by their offspring remained unknown. evaluate the relationship between preconception paternal body weight and their offspring’s birth weight in 1,810 Chinese mother-father-baby trios. Information on paternal and maternal preconception body weight and height was collected via a self-reported questionnaire. Birth weight was collected from medical records. Paternal preconception body weight was associated with offspring’s birth weight (*p trend=0*.*02*) after multivariable adjustment. Each standard deviation increment of paternal body mass index was associated with an additional 29.6 g increase of birth weight (95% confident interval: 5.7g, 53.5g). The association was more pronounced in male neonates, and neonates with overweight mothers, and with mothers who gained excessive gestational weight, compared to their counterparts (all p interaction<0.05). Sensitivity analyses showed similar pattern to that of the main analysis. Paternal preconception body weight was associated with birth weight of their offspring.

**Impact statement:** *What is already known on this subject?:* More efforts have previously been put on the maternal contribution to birth weight, however, it is uncertain whether paternal pre-conceptional body weight, an indicator for epigenetic information, might be inherited by their offspring.

*What do the results of this study add?:* In the current study that included 1,810 Chinese mother-father-baby trios, a small but significant association was observed between paternal preconception body weight and offspring’s birth weight (p trend=0.02).

*What are the implications of these findings for clinical practice and/or further research?:* Paternal epigenetic information of nutrition could be inherited by their offspring.

## Introduction

Most of the previous studies have been focused on birth weight dependent on maternal body weight. They found that both under- and overnutrition before conception was associated with unfavorable birth outcomes (Sebire *et al*., 2001, Black *et al*., 2013). These include the development of chronic diseases later in life - in particular obesity, diabetes, and cardiovascular diseases (Wahlqvist *et al*., 2015, Haire-Joshu and Tabak, 2016, Lowensohn *et al*., 2016). However, less is known about paternal contribution to weight status in the offspring. An animal study proved that male rats fed by a high-fat diet exhibited glucose intolerance. This nutritional influence on epigenetics could result in β-cell dysfunction of their female offspring, leading to similar phenomenon of glucose intolerance in their adulthood (Ng *et al*., 2010). Till now, there is a lack of human evidence to support this hypothesis (Abbasi, 2017). One cohort study reported that paternal body mass index (BMI) were associated with offspring BMI in both childhood and mid-adulthood, and the strength of the association did not reduce when offspring grew older (Cooper *et al*., 2010). However, potential environmental confounders were not controlled in these studies. In a small prospective study on this topic (n=372), paternal preconception BMI was associated with a non-significant increasing trend of birth weight (Moss and Harris, 2015). On the contrary, a cohort study including 2,947 singleton children in Netherland found that paternal BMI was associated with birthweight by univariate analysis, though no significant difference was evident following the adjustment of maternal factors (L’Abee *et al*., 2011). Another study performed in 889 Chinese mother-father-child triplet confirmed the association of paternal BMI with birthweight in sons, but not in daughters (Chen *et al*., 2012). Of note, many of these studies were limited by the absence of information on relevant factors during pregnancy (Kominiarek and Peaceman, 2017) and failure to exclude non-biological fathers in the analyses (Moss and Harris, 2015). A newly published review suggested that increased paternal BMI could negatively affect pregnancy and child health outcomes, while the association between parental BMI and birthweight cannot be established (Campbell and McPherson, 2019).

Therefore, we conducted the current study to examine the relationship between paternal preconception body weight (assessed by BMI) and their children’s birth weight in a large Chinese population. We hypothesized that paternal preconception body weight, which was considered as the epigenetic information, was associated with his children’s birth weight, independent of major maternal factors.

## Materials and Methods

### Study population

All the participants were recruited from the Department of Obstetrics, Ren Ji Hospital, School of Medicine, Shanghai Jiao Tong University, from 1^st^ January to 31^st^ December in 2015. The parent’s self-reported body weight and height before conception were obtained, as well as the newborns’ birth weight. The details of participant recruitment/exclusion were shown in **Supplemental Figure 1**. Finally, we included 1,810 trios of father, mother and their newborns in the current analysis. The study was approved by the Ethics Committee of Ren Ji Hospital, School of Medicine, Shanghai Jiao Tong University. This was a re-identified study based on a cohort study to evaluate maternal lipid profile and birth weight (Ref No. 2014QDQ10) and informed consent was given by both parent during their induction visit.

### Assessment of exposure (paternal/maternal body weight and height) and outcome (birth weight)

At the first prenatal checkup (∼15^th^ -16^th^ gestational weeks), we asked paternal and maternal preconception body weight and height in the past 6 months. Body mass index (BMI) was calculated as the body weight (in kilogram) divided by the height squared (in meters). Body weight status was used as both continuous and categorical variable. According to the Working Group of Obesity in China (WGOC) criteria for adults, participants’ weight status was classified as: ‘normal’ (BMI < 24.0 kg/m^2^), ‘overweight’ (24 kg/m^2^ ≤ BMI < 28.0 kg/m^2^), and ‘obese’ (BMI ≥ 28.0 kg/m^2^) (Zhou and Coorperative Meta-Analysis Group Of Working Group On Obesity In, 2002). As only 57 fathers (3.5%) had BMI categories as ‘underweight’, they were hence merged into ‘normal’ group for analysis. After birth delivery, we collected information on offspring’s birth weight from their medical records. Briefly, measurement of birth weight was completed within 1 hour of birth. This parameter was measured using a digital scale (Seca 757, Seca China, China) by a trained nurse in delivery room. Macrosomia is defined as birth weight is equal or more than 4000 g (Rossi *et al*., 2013).

### Assessment of covariates

Information on maternal age, education level, history of major chronic diseases (e.g., thyroid diseases, immune diseases, heart diseases, and psychiatric disorders), and family history of metabolic diseases (obese, pre-diabetes/diabetes, hypertension, dyslipidemia, nonalcoholic fatty liver diseases, and hyperuricemia/gout) were collected via a self-reported questionnaire by trained medical staffs. Data on parity, conception mode, and gestational weight gain (GWG) were obtained from medical records. All the mothers were categorized into three groups based on preconception BMI and GWG: inadequate GWC (below the lower cutoff point of IOM recommendation (Anon, 2009), optimal GWG (within the recommendation), and excessive GWG (above the upper cutoff point). The optimal GWG is 12.5-18.0 kg for underweight mother, 11.5-16.0 kg for normal weight mother, 7.0-11.5 kg for overweight mother, and 5.0-9.0 kg for obese mother. Data on hemoglobin, fasting plasma glucose, systolic blood pressure, diastolic blood pressure, plasma level of triglyceride, total cholesterol, low density lipoprotein cholesterol and high-density lipoprotein cholesterol at the first prenatal check-up were also obtained from medical record. Dyslipidemia was diagnosed if any of the following criterion was addressed: triglyceride≥1.7 mmol/L, total cholesterol≥5.72mmol/L, low density lipoprotein cholesterol≥3.4mmol/L, and high-density lipoprotein cholesterol<1.0 mmol/L)(Srikanth and Deedwania, 2016).

### Statistical analyses

All statistical analyses were conducted by SAS version 9.4 (SAS Institute, Inc, Cary, NC). Formal hypothesis testing was 2-sided with a significant level of 0.05.

We used PROC CORR to test parental BMI and offspring’s birthweight separately in boys and girls, adjusting for delivery gestational week (week), maternal age, GWG (kg), education (less than or equal to high school ***or*** college or above), parity (primipara ***or*** non-primipara), family history of diseases (‘yes’ ***or*** ‘no’), hemoglobin (g/L), systolic blood pressure (mmHg), diastolic blood pressure (mmHg), dyslipidemia (‘yes’ ***or*** ‘no’), and fasting plasma glucose (mmol/L) at the first prenatal check-up.

We used the **PROC GLM** to evaluate the relationship between parental body weight and offspring’s birthweight. Further, we used **PROC LOGISTIC** to evaluate the relationship between parental body weight and the risk of developing macrosomia. We considered many potential confounders for inclusion in each model. **Model 1** was adjusted for offspring’s sex and delivery gestational week. **Model 2** was adjusted for variables in ***model 1*** and further adjusted for maternal age, GWG (kg), preconception BMI (kg/m^2^), education (less than or equal to high school ***or*** college or above), family history of disease (‘yes’ ***or*** ‘no’) and parity (primipara ***or*** non-primipara). **Model 3** was adjusted for variables in ***model 2*** and further adjusted for hemoglobin (g/L), systolic blood pressure (mmHg), diastolic blood pressure (mmHg), dyslipidemia (‘yes’ ***or*** ‘no’), and fasting plasma glucose (mmol/L) at the first prenatal check-up.

Because offspring’s sex, maternal preconception BMI, and GWG might modify the association between paternal body weight and offspring’s birth weight, we tested relevant interaction and subgroup analyses based on these three variables, adjusting for aforementioned covariates.

To test the robustness of the results obtained from the main analysis, we conducted four sensitivity analyses: 1) we adjusted birth weight by delivery gestational week; 2) we adjusted birth weight by birth length as outcome; 3) we excluded mother aged 35 years old or more during pregnancy; and 4) excluding father whose BMI was less than 18.5 kg/m^2^.

## Results

There were 1,810 full-term healthy newborns (946 boys and 864 girls). The average birth weight was 3,415±395 g in boys and 3,314±376 g in girls. The average birth length was 50.1±1.4 cm and 49.9±1.0 cm for boys and girls respectively. The average gestational age at delivery was 39.0±1.1 weeks. Paternal body weight was associated with advanced maternal age, higher maternal preconception BMI and education level, higher prevalence of family history of metabolic diseases, and greater offspring’s birth weight (**Table 1**).

**Table 1.**
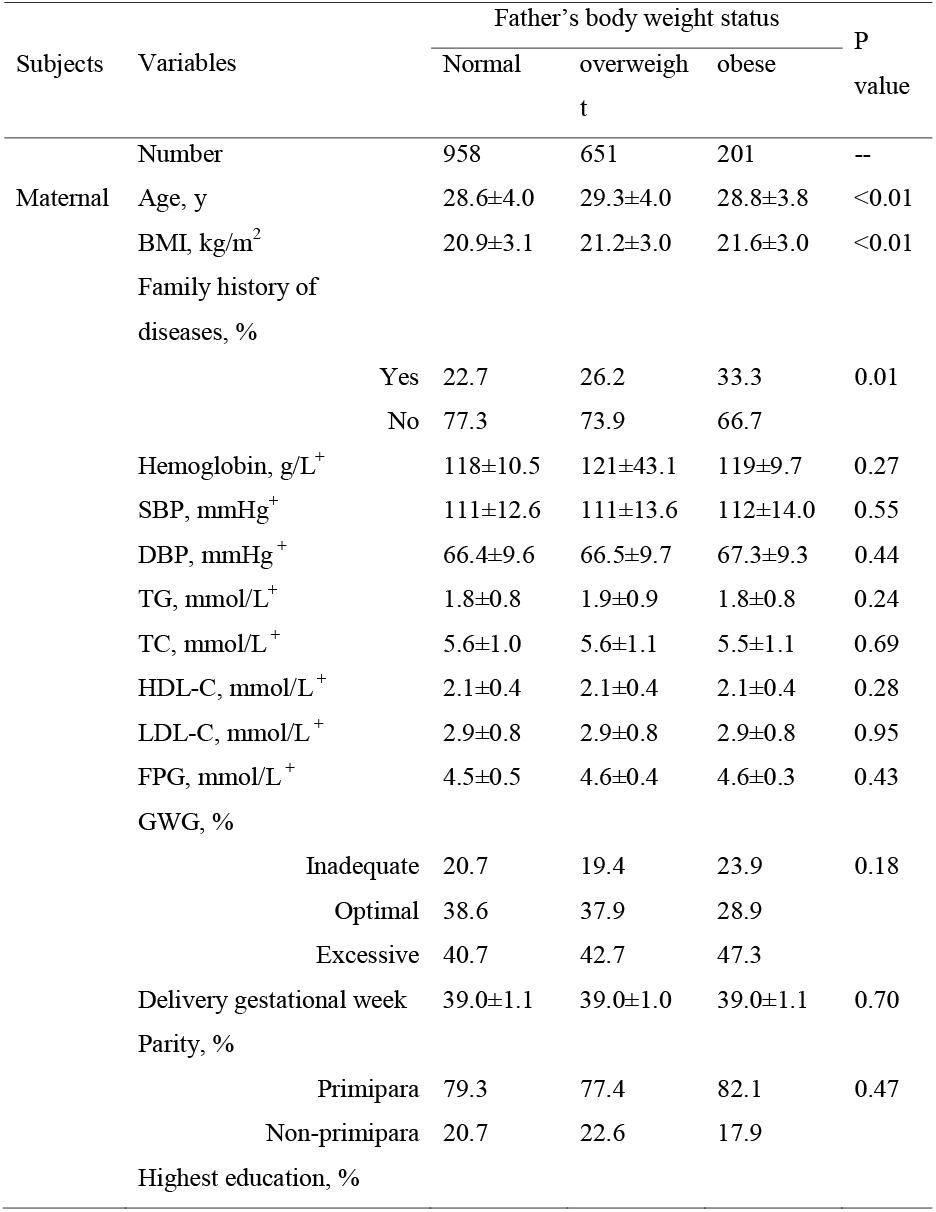
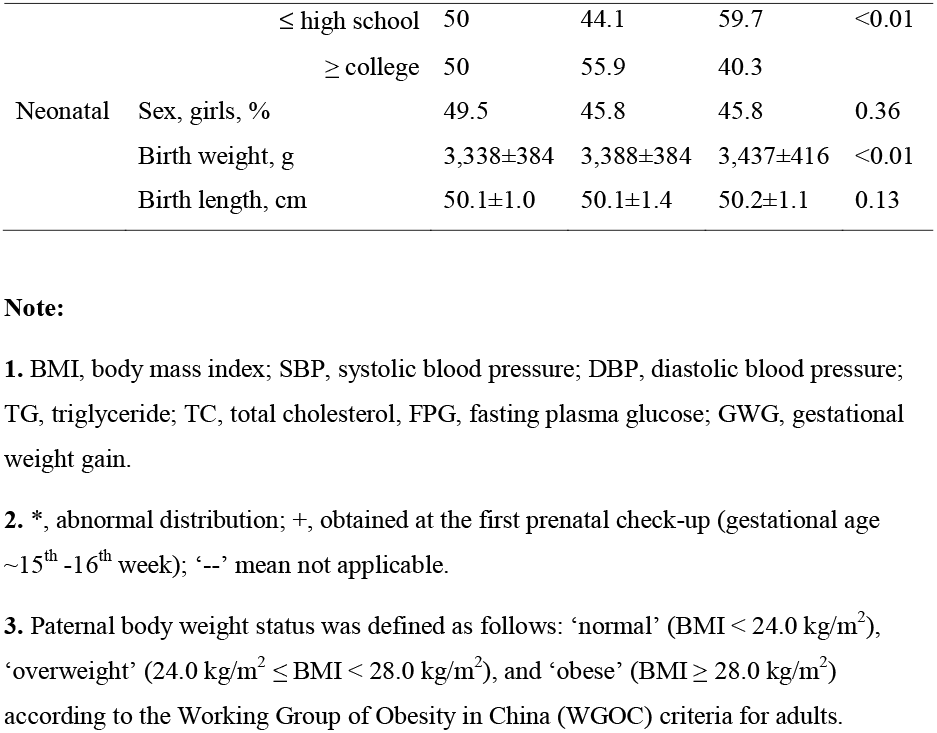
The maternal and neonatal baseline characteristics by paternal body weight status

Paternal preconception body weight was associated with offspring’s birth weight, after adjustment for potential confounders (**Table 2, model 2**). Further adjustment for hemoglobin, blood pressure, plasma lipids profiles, and fasting plasma glucose (mmol/L) at the first prenatal check-up attenuated the association; however, the difference between the two extreme groups remained significant. Each standard deviation (SD) increment of paternal BMI (≈3.27 kg/m^2^) was associated with an additional 29.6 g increase of birth weight [95% confident interval (CI): 5.7, 53.5 g, p trend=0.02] (**Table 2, model 3**). When considering paternal preconception BMI as a continuous variable, one-unit increase in BMI (1.0 kg/m^2^) was associated with a 9.6 g increase of offspring birth weight (95% CI: 2.3g, 17.0g, p=0.01). Paternal body weight was not associated with the risk of macrosomia (**Table 3, model 3**).

**Table 2.** Mean difference and 95% confidence intervals in offspring’s birth weight across three paternal groups in 1, 810 Chinese father-mother-baby trios

| Model | Father's body weight status |  |  | Per SD | P trend |
| --- | --- | --- | --- | --- | --- |
|  | Normal | Overweight | Obese |  |  |
| Number | 958 | 651 | 201 | / | / |
| Model 1 | **0 (Ref)** | 52.5 (16.5, 88.4) | 97.0 (42.1, 152) | 38.2 (21.6, 54.9) | <0.001 |
| Model 2 | **0 (Ref)** | 48.9 (13.8, 84.0) | 94.4 (40.7, 148) | 36.0 (19.6, 52.4) | <0.001 |
| Model 3 | **0 (Ref)** | 49.6 (-3.3, 102.5) | 87.3 (8.9, 165.6) | 35.5 (11.5, 59.4) | 0.004 |

**Note:**
**1.** Father's body weight status was defined as follows: 'normal' ( $\text{BMI} < 24.0 \text{ kg/m}^2$ ), 'overweight' ( $24.0 \text{ kg/m}^2 \leq \text{BMI} < 28.0 \text{ kg/m}^2$ ), and 'obese' ( $\text{BMI} \geq 28.0 \text{ kg/m}^2$ ) according to the Working Group of Obesity in China (WGOC) criteria for adults. '/' mean not applicable. Per SD for paternal $\text{BMI} \approx 3.27 \text{ kg/m}^2$ .
**2.** Dyslipidemia was diagnosed if any of the following was addressed: triglyceride $\geq 1.7 \text{ mmol/L}$ , total cholesterol $\geq 5.72 \text{ mmol/L}$ , low-density lipoprotein cholesterol $\geq 3.4 \text{ mmol/L}$ , high-density lipoprotein cholesterol $< 1.0 \text{ mmol/L}$ .
**3. Model 1** adjusted for offspring's sex and delivery gestational week (week).
**4. Model 2** adjusted for variables in **model 1** and further adjusted for maternal age, GWG (kg), BMI ( $\text{kg/m}^2$ ), education (less than or equal to high school *or* college or above), family history of metabolic diseases ('yes' *or* 'no') and parity (primipara *or* non-primipara).

**Table 3.** Adjusted Odd Ratios and 95% confidence intervals for risks of macrosomiaacross three paternal groups in 1, 810 Chinese father-mother-baby trios

| Model | Father's body weight status |  |  | Per SD | P trend |
| --- | --- | --- | --- | --- | --- |
|  | Normal | Overweight | Obese |  |  |
| Number | 958 | 651 | 201 | / | / |
| No. | 49 | 39 | 16 | / | / |
| Model 1 | **0 (Ref)** | 1.21 (0.78, 1.88) | 1.59 (0.88, 2.89) | 1.21 (1.001, 1.46) | 0.049 |
| Model 2 | **0 (Ref)** | 1.18 (0.76, 1.84) | 1.47 (0.8, 2.69) | 1.17 (0.97, 1.41) | 0.11 |
| Model 3 | **0 (Ref)** | 1.09 (0.57, 2.11) | 1.23 (0.5, 3.03) | 1.11 (0.84, 1.46) | 0.47 |

**Note:**
1. Paternal body weight status was defined as follows: ‘normal’ ( $\text{BMI} < 24.0 \text{ kg/m}^2$ ), ‘overweight’ ( $24.0 \text{ kg/m}^2 \leq \text{BMI} < 28.0 \text{ kg/m}^2$ ), and ‘obese’ ( $\text{BMI} \geq 28.0 \text{ kg/m}^2$ ) according to the Working Group of Obesity in China (WGOC) criteria for adults. Macrosomia was defined as birth weight was equal or greater than 4000 g.
2. ‘/’ mean not applicable. Per SD for paternal $\text{BMI} \approx 3.27 \text{ kg/m}^2$ .
3. Dyslipidemia was diagnosed if any of the following was addressed: triglyceride $\geq 1.7 \text{ mmol/L}$ , total cholesterol $\geq 5.72 \text{ mmol/L}$ , low-density lipoprotein cholesterol $\geq 3.4 \text{ mmol/L}$ , high-density lipoprotein cholesterol $< 1.0 \text{ mmol/L}$ .
4. **Model 1** adjusted for offspring’s sex and delivery gestational week (week).
5. **Model 2** adjusted for variables in **model 1** and further adjusted for maternal age, GWG (kg), BMI ( $\text{kg/m}^2$ ), education (less than or equal to high school *or* college or above), family history of metabolic diseases (‘yes’ *or* ‘no’) and parity (primipara *or* non-primipara).

The association between paternal preconception body weight and offspring’s birth weight appeared to be only present in male neonates, neonates with overweight mothers and mothers with excessive GWG, compared to their counterparts (p interaction <0.05 for all, **Figure 1**). Paternal preconception BMI was significantly associated with their sons’ birth weight while maternal preconception BMI was significantly associated with both sons and daughters’ birth weight (all p<0.01, **Table S 1**).

**Figure 1.**
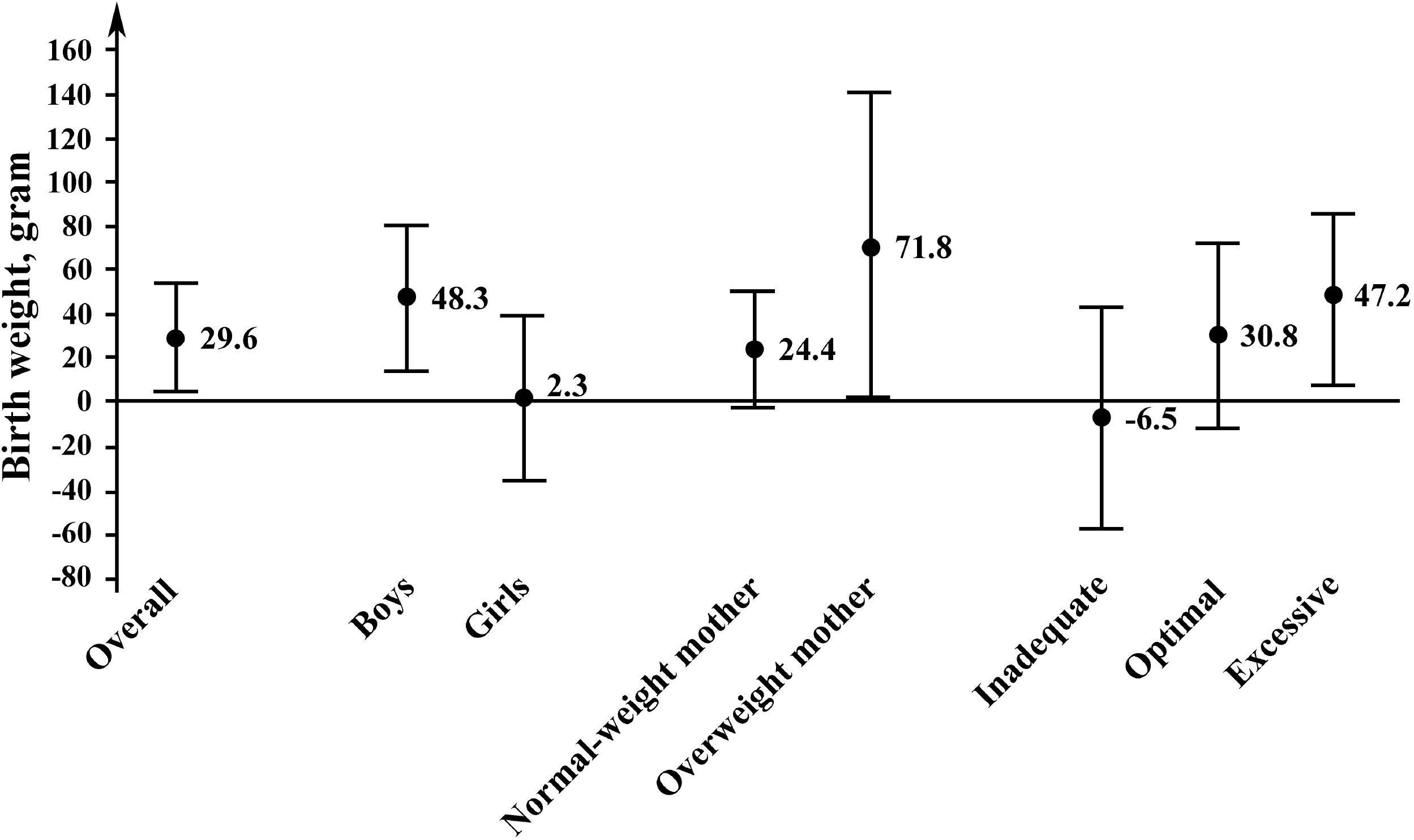
Mean difference and 95% confidence intervals in offspring’s birth weight associated with per SD increase of paternal preconception BMI in 1,810 Chinese mother-father-baby trios. **Note:** **1**. Interaction between paternal body weight status and offspring’s sex p value=0.04; interaction between maternal and paternal body weight status p value<0.001; interaction between paternal body weight status and gestational weight gain p value<0.001. **2**. Adjusted for offspring’s sex, delivery gestational week (week), maternal age, GWG (kg), BMI (kg/m^2^), education (less than or equal to high school, ***or*** college or above), parity (primipara ***or*** non-primipara), family history of diseases (‘yes’ ***or*** ‘no’), hemoglobin (g/L), systolic blood pressure (mmHg), diastolic blood pressure (mmHg), plasma level of triglyceride (mmol/L), total cholesterol (mmol/L), low density lipoprotein cholesterol (mmol/L), high density lipoprotein cholesterol (mmol/L), and fasting plasma glucose (mmol/L) at the first prenatal check-up (∼15^th^-16^th^ gestational week).

We found similar results in the sensitive analysis with gestational week adjusted weight, or birth length adjusted weight, as well as those excluding mothers aged 35 years or more, or fathers with underweight (BMI<18.5 kg/m^2^) during pregnancy **(Table S 2-5)**.

## Discussion

In this prospective study including 1,810 Chinese mother-father-baby trios, we found that paternal preconception body weight was associated with offspring’s birth weight, after adjustment for several potential confounders such as maternal preconception body weight. Our findings confirmed the theory that paternal BMI before pregnancy was possibly a risk factor for the gain of birth weight and thus might increase the future risk of developing obesity and related metabolic diseases. Hence, it is equally important to target paternal pre-pregnancy body weight as maternal body weight during preconception health advice.

Mothers have long been the focus of preconception care. Women of reproductive age are recommended to take folic acid, calcium, vitamin supplements, to taper off alcohol and smoking, reduce coffee consumption, and maintain healthy body weight (Abbasi, 2017). Although mothers take more responsibilities than fathers in conceiving a healthy baby, the influence of paternal pre-conception BMI on birth weight cannot be neglected (Campbell and McPherson, 2019). Paternal preconception environmental exposures also closely related to offspring’s birth weight (Shea and Little, 1997, Moss and Harris, 2015), and future health (Kaati *et al*., 2002, Yen *et al*., 2016) of their offspring. Detailed mechanism of transgenerational effects has not been elucidated yet, but was thought to be associated with spermatozoan coding RNAs (Krawetz, 2005), DNA methylation imprinted in sperm (Soubry *et al*., 2016), and identified features specific to sex chromosomes that related to epigenetic information (Pembrey *et al*., 2006, Navratilova *et al*., 2017). Consistently, we observed that preconception paternal body weight was associated with offspring’s birth weight. The potential role of paternal exposure on offspring heath status was supported by Shea and Little’s finding which had a group of fathers exposed to x-ray examinations within one year prior to conception. They found that the x-ray exposure had a marginally significant influence on birth weight, compared with those without it (3,358 g ***vs***. 3,437 g; p = 0.055). (Shea and Little, 1997). Likewise, in a longitudinal study (n=372) based on the National Longitudinal Study of Adolescent Health, preconception paternal BMI was associated with offspring’s birth weight (relative to father with normal weight, mean difference in birth weight was 35.6 g for overweight father, and 76.8 g for obese father) (Moss and Harris, 2015). The non-significant results of this study could be explained, at least in part, by its modest sample size (372 father-mother-infant trios) and crude estimation of biological relationship between father and children. Two previous cohort studies have been performed in 2,947 Netherland and 1,041 Australian children, respectively (L’Abee *et al*., 2011, Pomeroy *et al*., 2015). In accordance with the present study, no significant difference between paternal BMI and birthweight was evident. However, pre-term babies (<37 gestational weeks) were included in the analysis (L’Abee *et al*., 2011). Another possible explanation for the discrepancy between our results and previous studies might be due to the inclusion of multipara mothers in the analysis (L’Abee *et al*., 2011, Moss and Harris, 2015, Pomeroy *et al*., 2015).

In subgroup analysis, male neonates, neonates with overweight mothers, or with mothers who experienced excessive GWG were more likely to get greater birth weight, compared to their counterparts. Paternal epigenetic information seems to transfer to male offspring in sex-specific rule. In the current study, paternal BMI only correlated with their son’s, but not with their daughter’s birth weight. Our results were consistent with Chen’s study (Chen *et al*., 2012). In addition, Similar sex-specific transgenerational responses were also observed in Swedish population(Pembrey *et al*., 2006). Paternal smoking was associated with greater BMI at year 9 in sons, but not in daughters. Further, paternal grandfather’s food supply was only linked to the mortality of grandsons (Pembrey *et al*., 2006). A possible explanation for this is that paternal epigenetic information might leave marks on Y chromosome, hence is transferred only to male offspring (Pembrey *et al*., 2006, Navratilova *et al*., 2017). It is well recognized that maternal preconception obesity and higher gestational weight gain were associated with larger birth size (Strutz *et al*., 2012, Secher *et al*., 2014, Catalano and Shankar, 2017). Paternal effects on birth weight of their offspring were more pronounced only in neonates with overweight mother, or with mother who gained excessive weight in pregnancy. These results suggest that paternal effects could be modified by maternal factors during pregnancy.

Strengths of the current study included its longitudinal design, a large sample size, and precise assessment of the biological relationship between father and their children. Our study had several limitations. Firstly, both paternal and maternal body weight before pregnancy was self-reported, which might result in misclassification. However, self-reported weight is generally accurate and had high correlation with measured weight (Stunkard and Albaum, 1981, Leary *et al*., 2010). Secondly, residual confounding is of concern because paternal demographic information, such as age (Sharma *et al*., 2015), smoking (Han *et al*., 2015, Yen *et al*., 2016), and occupation (Friberg *et al*., 2015) were not collected and these missing data could be associated with offspring’s future health (Cheng *et al*., 2016). Finally, the macrosomia is considered as a more reliable parameter to predict the risk of future obesity and cardiovascular diseases compared to birth weight (Szostak-Wegierek, 2014, Woo Baidal *et al*., 2016). However, being limited to a lack of macrosomia cases (114/1810), the statistical power of the current study was low to explore the relationship between paternal body weight and risk of macrosomia, though positive association between the preconception paternal body weight and birth weight was confirmed.

In conclusion, paternal preconception body weight was associated with birth weight of their offspring. Future studies with detailed information on preconception paternal demographics are warranted to confirm our findings.

## Data Availability

The SAS code and data that support the findings of this study are available from the first author upon reasonable request (Renying Xu, email address)

## Data Sharing statement

The SAS code and data that support the findings of this study are available from the first author upon reasonable request (*Renying Xu*, email address).

## Supporting Information legends

**Supplemental Table 1**. The association between parental preconception BMI and their children’s birth weight in 1,810 Chinese mother-father-baby trios

**Supplemental Table 2**. Mean difference and 95% confidence intervals in offspring’s birth weight (g/week) adjusted by delivery gestational week, across three paternal groups in 1,810 Chinese mother-father-baby trios

**Supplemental Table 3**. Mean difference and 95% confidence intervals in offspring’s birth weight adjusted by birth length (g/cm), across three paternal groups in 1,810 Chinese mother-father-baby trios

**Supplemental Table 4**. Mean difference and 95% confidence intervals in offspring’s birth weight (g), across three paternal groups in 1,648 Chinese mother-father-baby trios, excluding women aged 35 years old or more

**Supplemental Table 5**. Mean difference and 95% confidence intervals in offspring’s birth weight (g), across three paternal groups in 1,753 Chinese mother-father-baby trios, excluding father with underweight

**Supplemental Figure 1**. Sample recruitment in the study.

