## Supplemental Materials for "Paternal body mass index before conception associated with offspring’s birth weight in Chinese population: a prospective study"

**Supplemental Table 1**. The association between parental preconception BMI and their children’s birth weight in 1,810 Chinese mother-father-baby trios

| Neonate | Father’s BMI* | Mother’s BMI^+^ |
| --- | --- | --- |
| Boys (n=954) | r=0.14  p=0.005 | r=0.18  p<0.001 |
| Girls (n=872) | r=-0.007  p=0.90 | r=0.26  p<0.001 |

**Note:**

**1.** BMI, body mass index.

**2**. Adjusted for delivery gestational week (week), maternal age, GWG (kg), education (less than or equal to high school ***or*** college or above), parity (primipara ***or*** non-primipara), family history of diseases (‘yes’ ***or*** ‘no’), hemoglobin (g/L), systolic blood pressure (mmHg), diastolic blood pressure (mmHg), plasma level of triglyceride (mmol/L), total cholesterol (mmol/L), low density lipoprotein cholesterol (mmol/L), high density lipoprotein cholesterol (mmol/L), and fasting plasma glucose (mmol/L) at the first prenatal check-up (~15^th^-16^th^ gestational week).

**3.** *, further controlled for mother’s BMI before conception. +, further controlled for father’s BMI before conception.

**Supplemental Table 2**. Mean difference and 95% confidence intervals in offspring’s birth weight (g/week) adjusted by delivery gestational week, across three paternal groups in 1,810 Chinese mother-father-baby trios

| Model | Father’s body weight status | | | Per SD | P trend |
| --- | --- | --- | --- | --- | --- |
|  | Normal | Overweight | Obese |  |  |
| Number | 958 | 651 | 201 | / | / |
| Model 1 | **0 (Ref)** | 1.31 (0.38, 2.24) | 2.48 (1.07, 3.9) | 0.97 (0.54, 1.4) | <0.001 |
| Model 2 | **0 (Ref)** | 1.23 (0.33, 2.14) | 2.44 (1.05, 3.82) | 0.92 (0.49, 1.34) | <0.001 |
| Model 3 | **0 (Ref)** | 1.35 (-0.04, 2.73) | 2.33 (0.28, 4.38) | 0.94 (0.32, 1.57) | 0.003 |

**Note:**

**1.** Parental body weight status was defined as follows: ‘normal’ (BMI < 24.0 kg/m^2^), ‘overweight’ (24.0 kg/m^2^ ≤ BMI < 28.0 kg/m^2^), and ‘obese’ (BMI ≥ 28.0 kg/m^2^) according to the Working Group of Obesity in China (WGOC) criteria for adults. ‘/’ mean not applicable.

**2. Dyslipidemia was diagnosed if any of the following was confirmed:** triglyceride≥1.7 mmol/L; total cholesterol≥5.72mmol/L; low-density lipoprotein cholesterol≥3.4mmol/L; high-density lipoprotein cholesterol<1.0 mmol/L.

**3. Model 1** adjusted for offspring’s sex.

**4.** **Model 2** adjusted for variables in model 1 and further adjusted for maternal age, GWG (kg), BMI (kg/m^2^), education (less than or equal to high school ***or*** college or above), family history of metabolic diseases (‘yes’ ***or*** ‘no’) and parity (primipara ***or*** non-primipara).

**5. Model 3** adjusted for variables in model 2 and further adjusted for hemoglobin (g/L), systolic blood pressure (mmHg), diastolic blood pressure (mmHg), dyslipidemia (‘yes’, ***or*** ‘no’), and fasting plasma glucose (mmol/L) at the first prenatal check-up (~15^th^-16^th^ gestational week).

**Supplemental Table 3**. Mean difference and 95% confidence intervals in offspring’s birth weight adjusted by birth length (g/cm), across three paternal groups in 1,810 Chinese mother-father-baby trios

| Model | Father’s body weight status | | | Per SD | P trend |
| --- | --- | --- | --- | --- | --- |
|  | Normal | Overweight | Obese |  |  |
| Number | 958 | 651 | 201 | / | / |
| Model 1 | **0 (Ref)** | 0.93 (0.28, 1.59) | 1.67 (0.67, 2.67) | 0.67 (0.37, 0.98) | <0.001 |
| Model 2 | **0 (Ref)** | 0.87 (0.23, 1.51) | 1.60 (0.63, 2.58) | 0.63 (0.33, 0.92) | <0.001 |
| Model 3 | **0 (Ref)** | 0.85 (-0.08, 1.79) | 1.39 (0.004, 2.77) | 0.59 (0.17, 1.02) | 0.006 |

**Note:**

**1.** Parental body weight status was defined as follows: ‘normal’ (BMI < 24.0 kg/m^2^), ‘overweight’ (24.0 kg/m^2^ ≤ BMI < 28.0 kg/m^2^), and ‘obese’ (BMI ≥ 28.0 kg/m^2^) according to the Working Group of Obesity in China (WGOC) criteria for adults. ‘/’ mean not applicable.

**2. Dyslipidemia was diagnosed if any of the following was confirmed:** triglyceride≥1.7 mmol/L; total cholesterol≥5.72mmol/L; low-density lipoprotein cholesterol≥3.4mmol/L; high-density lipoprotein cholesterol<1.0 mmol/L.

**3.** **Model 1** adjusted for offspring’s sex and delivery gestational week (week).

**4.** **Model 2** adjusted for variables in model 1 and further adjusted for maternal age, GWG (kg), BMI (kg/m^2^), education (less than or equal to high school ***or*** college or above), family history of metabolic diseases (‘yes’ ***or*** ‘no’) and parity (primipara ***or*** non-primipara).

**5. Model 3** adjusted for variables in model 2 and further adjusted for hemoglobin (g/L), systolic blood pressure (mmHg), diastolic blood pressure (mmHg), dyslipidemia (‘yes’, ***or*** ‘no’), and fasting plasma glucose (mmol/L) at the first prenatal check-up (~15^th^-16^th^ gestational week).

**Supplemental Table 4**. Mean difference and 95% confidence intervals in offspring’s birth weight (g), across three paternal groups in 1,648 Chinese mother-father-baby trios, excluding women aged 35 years old or more

| Model | Father’s body weight status | | | Per SD | P trend |
| --- | --- | --- | --- | --- | --- |
|  | Normal | Overweight | Obese |  |  |
| Number | 879 | 582 | 187 | / | / |
| Model 1 | **0 (Ref)** | 60.9 (23.2, 98.6) | 111 (54.2, 168) | 43.3 (26.1, 60.4) | <0.001 |
| Model 2 | **0 (Ref)** | 57.6 (21.0, 94.1) | 109 (53.8, 164.2) | 41.5 (24.8, 58.3) | <0.001 |
| Model 3 | **0 (Ref)** | 53.7 (-1.5, 108.6) | 94.5 (15.0, 174) | 39.0 (14.6, 63.3) | 0.001 |

**Note:**

**1.** Parental body weight status was defined as follows: ‘normal’ (BMI < 24.0 kg/m^2^), ‘overweight’ (24.0 kg/m^2^ ≤ BMI < 28.0 kg/m^2^), and ‘obese’ (BMI ≥ 28.0 kg/m^2^) according to the Working Group of Obesity in China (WGOC) criteria for adults. ‘/’ mean not applicable.

**2. Dyslipidemia was diagnosed if any of the following was confirmed:** triglyceride≥1.7 mmol/L; total cholesterol≥5.72mmol/L; low-density lipoprotein cholesterol≥3.4mmol/L; high-density lipoprotein cholesterol<1.0 mmol/L.

**3.** **Model 1** adjusted for offspring’s sex and delivery gestational week (week).

**4.** **Model 2** adjusted for variables in model 1 and further adjusted for maternal age, GWG (kg), BMI (kg/m^2^), education (less than or equal to high school ***or*** college or above), family history of metabolic diseases (‘yes’ ***or*** ‘no’) and parity (primipara ***or*** non-primipara).

**5. Model 3** adjusted for variables in model 2 and further adjusted for hemoglobin (g/L), systolic blood pressure (mmHg), diastolic blood pressure (mmHg), dyslipidemia (‘yes’, ***or*** ‘no’), and fasting plasma glucose (mmol/L) at the first prenatal check-up (~15^th^-16^th^ gestational week).

**Supplemental Table 5**. Mean difference and 95% confidence intervals in offspring’s birth weight (g), across three paternal groups in 1,753 Chinese mother-father-baby trios, excluding father with underweight

| Model | Father’s body weight status | | | Per SD | P trend |
| --- | --- | --- | --- | --- | --- |
|  | Normal | Overweight | Obese |  |  |
| Number | 901 | 651 | 201 | / | / |
| Model 1 | **0 (Ref)** | 50.8 (14.1, 87.4) | 95.3 (39.8, 150.9) | 41.7 (24.1, 59.4) | <0.001 |
| Model 2 | **0 (Ref)** | 49.1 (13.4, 84.8) | 94.4 (40.2, 148.6) | 40.3 (23.0, 57.6) | <0.001 |
| Model 3 | **0 (Ref)** | 50.6 (-3.0, 104.3) | 88.2 (9.0, 167.4) | 37.5 (12.5, 62.5) | 0.003 |

**Note:**

**1.** Parental body weight status was defined as follows: ‘normal’ (BMI < 24.0 kg/m^2^), ‘overweight’ (24.0 kg/m^2^ ≤ BMI < 28.0 kg/m^2^), and ‘obese’ (BMI ≥ 28.0 kg/m^2^) according to the Working Group of Obesity in China (WGOC) criteria for adults. ‘/’ mean not applicable.

**2.** Underweight was diagnosed if paternal BMI was less than 18.5 kg/m^2^.

**3. Dyslipidemia was diagnosed if any of the following was confirmed:** triglyceride≥1.7 mmol/L; total cholesterol≥5.72mmol/L; low-density lipoprotein cholesterol≥3.4mmol/L; high-density lipoprotein cholesterol<1.0 mmol/L.

**4.** **Model 1** adjusted for offspring’s sex and delivery gestational week (week).

**5.** **Model 2** adjusted for variables in model 1 and further adjusted for maternal age, GWG (kg), BMI (kg/m^2^), education (less than or equal to high school ***or*** college or above), family history of metabolic diseases (‘yes’ ***or*** ‘no’) and parity (primipara ***or*** non-primipara).

**6. Model 3** adjusted for variables in model 2 and further adjusted for hemoglobin (g/L), systolic blood pressure (mmHg), diastolic blood pressure (mmHg), dyslipidemia (‘yes’, ***or*** ‘no’), and fasting plasma glucose (mmol/L) at the first prenatal check-up (~15^th^-16^th^ gestational week).

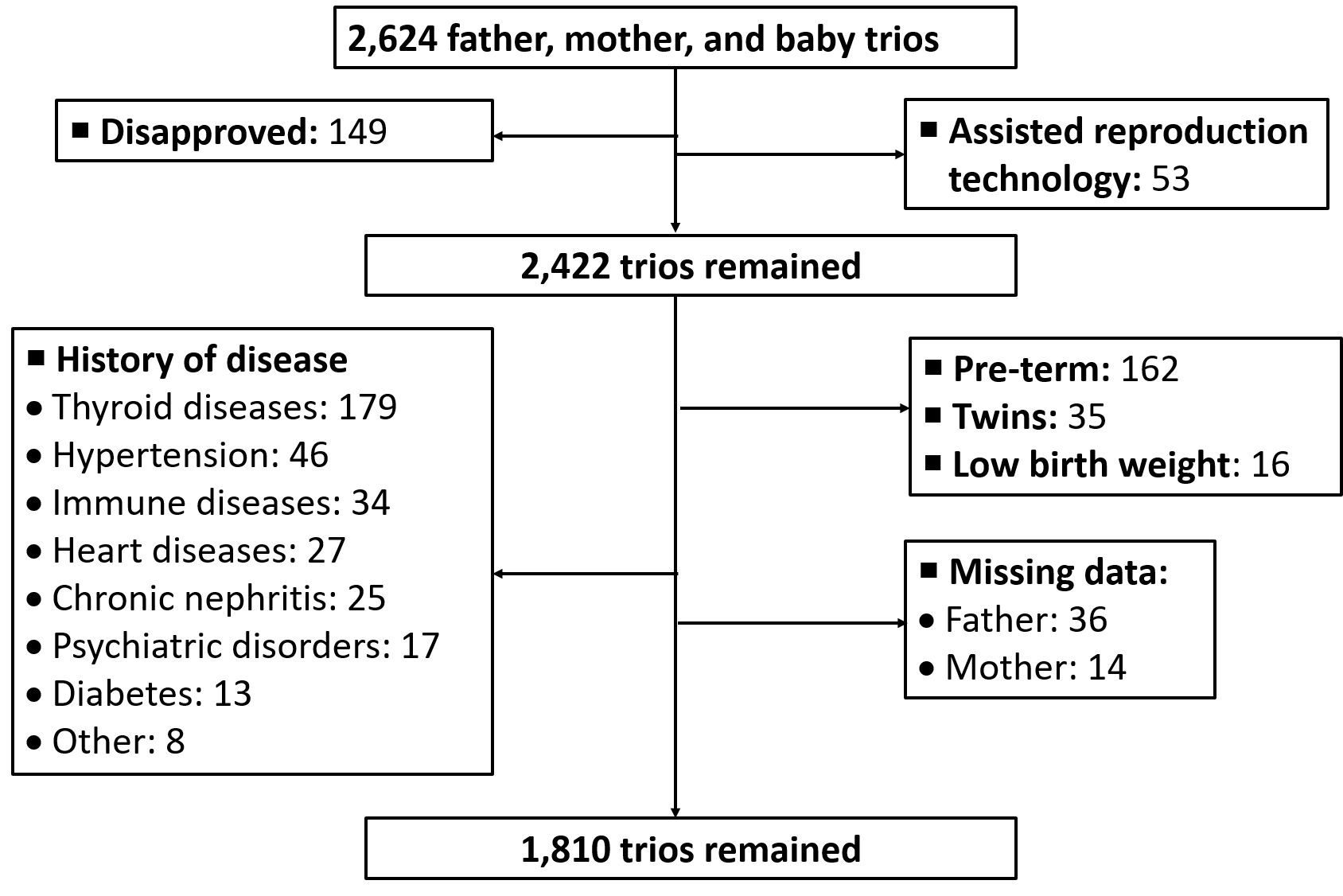

**Supplemental Figure 1**. Sample recruitment in the study. Thyroid diseases included 121 patients with hypothyroidism, 29 with hyperthyroidism, 15 with Hashimoto's thyroiditis, and 14 with thyroid adenoma. Immune diseases included 23 patients with systematic lupus erythematosus (SLE), 5 patients with Sjogren's syndrome, 3 patients with ankylosing spondylitis (AS), 2 patients with rheumatoid arthritis, and 1 patient with mixed connective tissue disease. Heart diseases included 23 patients with congenital disease and 4 with rheumatic heart disease. Psychiatric disorders included 11 patients with depression and 6 patients with epilepsy. Other diseases included 5 patients with syphilis infection, 2 with thalassemia, and 1 with ulcerative colitis. Low birth weight mean birth weight was less than 2500 grams.
